# Do Not Resuscitate practices in ICU. Descriptive study

**DOI:** 10.1101/2025.05.31.25328712

**Authors:** Mohammed A. Al-Odat, Waleed Aletreby, Hend M. Hamido

## Abstract

**Background:** Do Not Resuscitate (DNR) orders represent one of the most ethically complex decisions in the intensive care unit (ICU). Despite their significance, the implementation and communication of DNR decisions remain inconsistent, particularly in regions like the Middle East where data are limited.

**Objective:** To describe current DNR practices in a tertiary ICU in Saudi Arabia, focusing on the frequency, timing, context of issuance, and family involvement in DNR decisions.

**Methods:** This retrospective descriptive study included adult patients (16 years or older) who were discharged from a large Ministry of Health ICU in central Saudi Arabia between January 1 and March 31, 2025, and had a documented DNR order. Demographic data, clinical characteristics, and DNR decision details were extracted from electronic medical records.

**Results:** Of 889 ICU discharges, 168 patients died, and 77 (45.8%) had a DNR order. The average age of DNR patients was 51 (22) years; 62.3% were male. Common diagnoses included sepsis/septic shock (28.6%), malignancies (14.3%), and ischemic stroke (13%). Only 24.7% of DNR orders were issued within 48 hours of ICU admission, while 62.3% followed a successful cardiopulmonary resuscitation (CPR). Family involvement in the DNR decision was documented in only 22.1% of cases. All secondary outcomes late DNR issuance, post-CPR DNR decisions, and limited family involvement were statistically significant (p < 0.05).

**Conclusion:** DNR orders in this ICU were often delayed, issued reactively after CPR, and made without informing or involving families. These findings highlight the need for timely, proactive, and communicative end-of-life planning. Institutional policies and clinician training are essential to promote ethically sound and patient-centered DNR practices.

## Introduction

The intensive care unit (ICU) is a complex clinical environment where critically ill patients receive advanced life-sustaining interventions (1). Among the most ethically sensitive and clinically challenging decisions in the ICU is the establishment of Do Not Resuscitate (DNR) orders (1, 2). A DNR order indicates that, in the event of cardiac or respiratory arrest, cardiopulmonary resuscitation (CPR) should not be initiated. These decisions are often made in the context of poor prognosis, terminal illness, or patient wishes, with the aim of avoiding non-beneficial or potentially harmful interventions (3, 4).

DNR practices vary significantly across countries, institutions, and even among individual healthcare providers (6, 7). Factors influencing DNR decision-making include patient autonomy, cultural and religious values, legal frameworks, institutional policies, and the clinical judgment of physicians (7-9). Despite the ethical and clinical importance of these decisions, research shows that DNR orders are often delayed, inconsistently applied, or poorly communicated, leading to confusion among care teams and distress for patients and families (1).

In Middle Eastern countries, including Saudi Arabia, studies on DNR practices remain limited. There is a need to better understand how DNR decisions are made, documented, and perceived in ICU settings. By exploring these practices, we can identify gaps in communication, policy implementation, and education, thereby contributing to more patient-centered and ethically sound end-of-life care.

This study aims to describe current DNR practices in the ICU, including the frequency of DNR orders, the timing of DNR discussions, their relation with incidence of cardiac arrest, and the role of patients and families in these decisions. Understanding these aspects is essential for improving end-of-life care and supporting healthcare providers in making ethically appropriate decisions.

## Method and subjects

### Timeframe and setting

This study was conducted in the ICU of the largest Ministry of Health hospital in the central region of Saudi Arabia. The ICU includes 110 beds, divided into medical, surgical, respiratory, neuro-critical, trauma, burn, and obstetrics and gynecology units. All ICU beds are fully equipped with invasive and non-invasive monitoring and ventilation technologies. The ICU is run by intensivists round the clock, with a one is to one nursing to patient ratio.

The study utilizes retrospectively collected data of ICU patients between January 1^st^, and March 31^st^, 2025. Recorded data included demographic variables such as age and sex, diagnostic broad category as: medical, surgical, trauma, or burn, in addition to the actual diagnosis, along with Acute Physiologic Assessment and Chronic Health Evaluation (APACHE) four score, calculated using worst values of the first 24 hours in ICU (10). This study was approved by the local institutional review board with waiver of consent, in view of its retrospective design.

### Inclusion and exclusion criteria

We included in this study all patients discharged from the ICU between January and March 2025, who were 16 years old or more, stayed in the ICU at least 24 hours, and had a DNR form completed in their electronic health record. Patients who were discharged from the ICU without being labeled as DNR were excluded, regardless of whether they were alive or dead.

### Study objectives

The primary objective was to report the number and percentage of patients discharged from the ICU with a DNR order out of all discharged from ICU. Secondary objectives included describing and comparing DNR orders with regards to timing (early versus late), an early DNR order is issued within 48 hours of admission to ICU, DNR orders issued later on are considered late.

Whether the DNR order was issued following an episode of cardio-pulmonary arrest, and a successful cardio-pulmonary resuscitation (CPR), and whether informing the family of the DNR order is documented.

### Statistical plan

Continuous variables were summarized as mean and standard deviation, whereas discrete variables were presented as frequency (count) and percentage. Comparisons of DNR timing, following CPR, and informed families took place by Chi Square test. P values less than 0.05 were considered statistically significant. There were no correction for multiple testing, hence, results of statistical comparisons should be interpreted cautiously.

### Results

During the study period there were 889 patients discharged from the ICU, of which 168 patients died (including DNR cases), which represents a mortality rate of 18.9%, out of the 168 fatalities, 77 patients had a confirmed DNR order in their medical records. This represents 45.8% of all fatalities.

The average age of DNR patents was 51 ± 22 years, including 29 females (37.7%) and 48 males (62.3%). DNR patients had an average length of stay (LOS) of 20 ± 18.5 days in the ICU. All DNR patients died in the ICU (Table 1).

**Table 1:** Demographics of DNR patients:

| Variable (n = 77) | Mean ± SD / n (%) |
| --- | --- |
| Age (years) | $51 \pm 22$ |
| Sex: Males | 48 (62.3%) |
| LOS (days) | $20 \pm 18.5$ |

The most common three diagnoses among DNR patients was sepsis or septic shock (28.6%), followed by malignancies (solid or hematological) (14.3%), then ischemic stroke (13%) (Figure 1).

**Figure 1:**
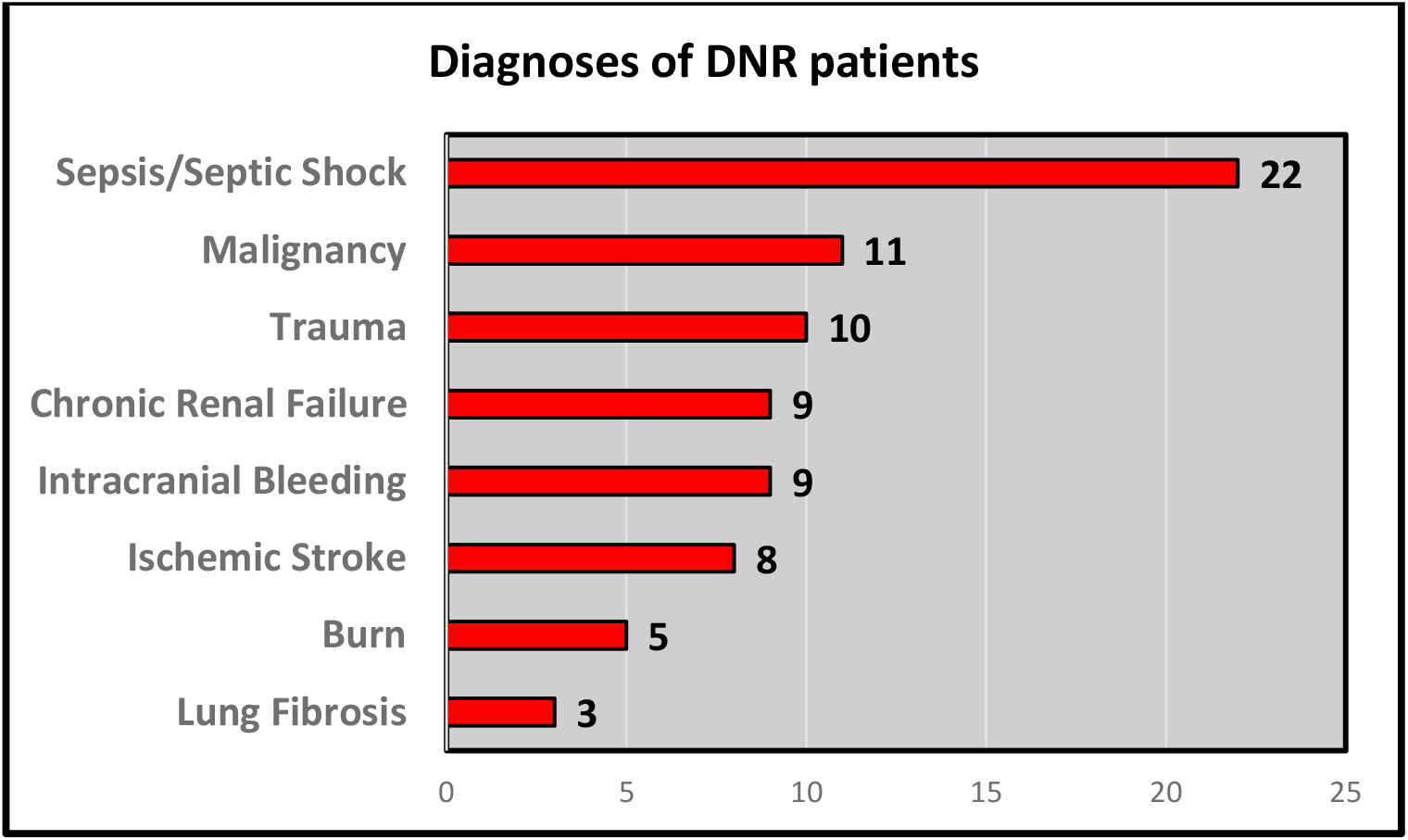
Diagnoses of DNR patients:

Among the 77 DNR patients, only 19 (24.7%) DNR orders were issued within 48 hours of ICU admission, whereas, 58 (75.3%) were considered late. The DNR order was issued following an episode of cardiac arrest and successful CPR in 48 (62.3%) cases, while only in 29 (37.7%) cases prior to cardiac arrest. The family of the patient was involved in the DNR decision or at least informed of it in only 17 (22.1%) cases, while, in 60 (77.9%) cases they were not involved in taking the decision nor informed. The three comparisons of the secondary outcomes were statistically significant, with p values of < 0.001, 0.002, and < 0.001 respectively (Table 2).

**Table 2:** Secondary outcomes:

| Variable (n = 77) | Yes: n, (%) | No: n (%) | Difference (95% CI); p value |
| --- | --- | --- | --- |
| **Early DNR (within 48 hrs. of ICU admission)** | 19 (24.7%) | 58 (75.3%) | -50.6 (-64.3 to -37);<br>< 0.001 |
| **DNR order following CPR** | 48 (62.3%) | 29 (37.7%) | 24.7 (9.4 to 40);<br>0.002 |
| **Family informed / involved in DNR decision** | 17 (22.1%) | 60 (77.9%) | -55.8 (-68.9 to -42.7),<br>< 0.001 |

## Discussion

This study provides a descriptive overview of DNR practices in a tertiary ICU setting, highlighting critical trends in timing, clinical context, and family involvement. Out of the 168 ICU deaths during the study period, 77 (45.8%) had documented DNR orders. This DNR rate aligns with the lower to moderate range reported in literature, which varies widely depending on cultural, institutional, and legal factors, with rates ranging from 30% to over 65% in end-of-life cases in some Western settings (11-13).

The average age of patients with DNR orders was 51 years, which is relatively young compared to similar studies. This may reflect differences in patient demographics or ICU admission criteria in our setting. The predominance of male patients and the average ICU length of stay of 20 days suggest that these patients had prolonged and likely complex clinical courses. The most common diagnoses among DNR patients (sepsis/septic shock, malignancies, and ischemic stroke) are consistent with conditions often associated with high ICU mortality and poor prognosis (12, 14).

A particularly noteworthy finding was the timing of DNR orders. Only 24.7% of DNR orders were issued within the first 48 hours of ICU admission, with the majority (75.3%) considered late, similar to our previously reported percentages (1), and similar to that reported by others (15). This delay may indicate a reluctance or systemic barrier to initiating end-of-life discussions early in the patient’s trajectory, possibly due to prognostic uncertainty, lack of clear protocols, or discomfort among healthcare providers in addressing such decisions early.

Additionally, 62.3% of DNR decisions were made only after a cardiac arrest and successful CPR, highlighting a reactive rather than proactive approach. Issuing DNR orders post-CPR can undermine the ethical principle of avoiding potentially non-beneficial interventions and may lead to unnecessary suffering. Best practices advocate for anticipatory decision-making based on prognosis, patient values, and clinical judgment (3, 16).

Equally concerning is the limited involvement of families, with only 22.1% of cases including family in the decision-making process or even informing them. This raises serious ethical considerations, as family-centered care is a cornerstone of ICU practice. Failure to engage families not only impacts trust and satisfaction but may also conflict with patients’ previously expressed wishes, especially in settings where advance directives are uncommon or legally unrecognized. It is true that in Saudi Arabia the consent of the family is not required to issue a DNR order, as it is considered a pure medical decision (17, 18), however, it is highly recommended to involve the family in making such a decision, to be aware of realistic expectations for their relatives (19).

All three secondary outcomes (late DNR, post-CPR DNR, and lack of family involvement) were statistically significant, underscoring systemic patterns rather than random occurrences. These findings suggest a need for institutional policies to guide timely and ethically sound DNR practices.

### Limitations

This study was conducted at a single center, which may limit the generalizability of results. Additionally, data were based on documentation in medical records, which may not fully capture informal discussions or undocumented decision-making processes. The retrospective nature of the study also limits our ability to explore clinician reasoning or patient preferences.

### Implications and Future Directions

Our findings highlight an urgent need to improve early goals-of-care discussions in the ICU. Interventions such as implementing standardized DNR protocols, training staff in communication skills, and promoting early family meetings could enhance both the timing and quality of DNR decision-making. Future research should explore barriers to timely DNR orders and evaluate the impact of structured communication strategies on patient and family outcomes.

## Conclusion

This study highlights significant delays in the initiation of DNR orders, a high frequency of reactive decisions following CPR, and limited family involvement in end-of-life discussions in our ICU setting. These findings suggest opportunities for improving the quality of end-of-life care through earlier and more proactive DNR decision-making and better communication with families. Implementing structured protocols and enhancing clinician training in goals-of-care conversations may promote more patient-centered and ethically sound practices in the ICU.

## Data Availability

All data produced in the present study are available upon reasonable request to the authors

